# Associations between ZIP code-level alcohol outlet density and binge drinking among people who inject drugs in 22 US Metropolitan Areas

**DOI:** 10.1101/2025.06.23.25329062

**Authors:** Snigdha R. Peddireddy, Stephanie Beane, Courtney Yarbrough, Umedjon Ibragimov, Janet R. Cummings, Danielle F. Haley, Sabriya L. Linton, Hannah L.F. Cooper

**Affiliations:** Department of Behavioral, Social, and Health Education Sciences, Rollins School of Public Health, Emory University, 1518 Clifton Road NE, Atlanta, GA 30322, USA; Department of Health Policy and Management, Rollins School of Public Health, Emory University, 1518 Clifton Road NE, Atlanta, GA 30322, USA; Department of Community Health Sciences, School of Public Health, Boston University, 715 Albany Street, Boston, MA 02118, USA; Department of Mental Health, Bloomberg School of Public Health, Johns Hopkins University, 615 N Wolfe Street, Baltimore, MD 21205, USA

**Keywords:** alcohol outlet density, binge drinking, injection drug use, equity, prevention, SDOH

## Abstract

**Background:** Although alcohol outlet density (AOD) is associated with drinking behaviors in the general population, little evidence exists about this relationship among people who inject drugs (PWID). Establishing this connection is particularly important, as alcohol use exacerbates the risk of opioid overdose. This study investigated 1) the association between ZIP code-level AOD and binge drinking among PWID who use opioids, and 2) the potential moderating effect of race/ethnicity on the AOD-binge drinking relationship.

**Methods:** This analysis linked 2018 National HIV Behavioral Surveillance (NHBS) data and 2016 ZIP code-level AOD data from the ZIP Code Business Pattern survey. Hierarchical generalized linear models quantified the association between AOD and binge drinking, overall and by race/ethnicity.

**Results:** Of 9,660 PWID, 27% reported recent binge drinking. For Hispanic/Latinx and White PWID, increases in AOD were associated with a 1% and 3% increase in the odds of binge drinking, respectively (p < 0.05); AOD was unrelated to binge drinking among Black PWID. However, when AOD was set to the median, binge drinking probability was highest among Black PWID (PP: 0.28, 95% CI: 0.24-0.33).

**Conclusions:** Reducing AOD may decrease binge drinking among White and Latinx PWID. Our results suggest, however, that this strategy alone may be not be effective among Black PWID. Holistic strategies targeting structural determinants of alcohol and opioid co-use among Black PWID are needed to address inequities in drug-related harms.

## 1. INTRODUCTION

Amidst the ongoing US opioid overdose crisis, overdose risk is particularly elevated among individuals who simultaneously consume alcohol and opioids. Opioids cause overdoses by suppressing respiration, and consuming multiple alcoholic drinks in one occasion (i.e., binge drinking) may further escalate overdose risk by compounding these suppressive effects (van der Schrier et al., 2017). The magnitude of alcohol’s contribution to the opioid overdose epidemic is significant: as of 2017, approximately 15% of heroin and fentanyl overdose deaths across the United States involved alcohol (Tori et al., 2020). Also, state-level binge drinking rates were correlated with alcohol’s involvement in opioid overdoses.

Evidence from the general population indicates that patterns of binge drinking are significantly shaped by spatial access to alcohol outlets. Studies have established a strong positive association between alcohol outlet density (AOD) and multiple alcohol-related outcomes, including the average volume consumed, frequency of drinking, and binge drinking (Bryden et al., 2012; Campbell et al., 2009; Popova et al., 2009; Xuan et al., 2015). AOD is also associated with overdose: in Baltimore City, each additional off-premise outlet (i.e., an outlet where alcohol is sold for consumption off-premise, including liquor and convenience stores) was associated with a 16.6% increase in the drug overdose rate among residents within a census block group (Nesoff et al., 2021). These and other findings have formed the evidence base for policies that support the health of the general population by limiting AOD, promoted by the Community Preventive Services Task Force, Substance Abuse and Mental Health Services Administration, and other public health bodies (Campbell et al., 2009; Robert Wood Johnson Foundation & University of Wisconsin Population Health Institute, n.d.; Substance Abuse and Mental Health Services Administration, 2022). The regulation of AOD may occur through zoning ordinances that delineate specific areas where alcohol sales are prohibited or that set minimum distance requirements between outlets. Regulation may also include licensing laws that cap the number of licenses available per locality or that set population-based quotas for alcohol licenses (Sacks et al., 2020).

While researchers have comprehensively explored relationships between AOD and an array of health outcomes among the general population, no studies have investigated the link between AOD and binge drinking among people who use non-prescribed opioids. Generating this evidence is essential due to this population’s elevated risk of overdose. Moreover, people who *inject* opioids are at an especially amplified risk for overdose: injecting delivers drugs into the bloodstream far more rapidly and with greater potency (Mathers et al., 2013). Binge drinking also exacerbates vulnerability to multiple other injection-related harms, including hepatitis C and HIV (Arasteh et al., 2008; Irvin et al., 2019).

Research has also neglected the extent to which (1) the relationship between AOD and binge drinking varies by race/ethnicity among people who inject drugs (PWID), and (2) AOD is racialized among PWID. The racialization of AOD is well-established for the general population: across major urban areas in the US, the density of off-premise outlets is often higher in neighborhoods inhabited predominantly by non-Hispanic/Latinx Black or Hispanic/Latinx residents compared to neighborhoods with mostly non-Hispanic/Latinx White residents (Berke et al., 2010; Fliss et al., 2021; LaVeist & Wallace, 2000; Lee et al., 2020; Romley et al., 2007). This phenomenon is explained in part by a history of racist policies (e.g., redlining) and land use practices that allowed for outlet proliferation in the neighborhoods where these communities live (Fliss et al., 2021; Haley, Jardine, et al., 2023; LaVeist & Wallace, 2000; Lee et al., 2020; Romley et al., 2007; Trangenstein et al., 2020). The racialization of alcohol risk environments may intersect with rapidly increasing rates of binge drinking (Barbosa et al., 2021) to exacerbate disproportionately higher overdose rates among non-Hispanic/Latinx Black and American Indian/Alaska Native individuals (Friedman & Hansen, 2022; Han et al., 2022; Kariisa et al., 2022). To date, no studies have explored whether the relationship between AOD and drinking behaviors varies by race/ethnicity, although Xuan et al. (2015) modeled that the implementation of stronger state-level alcohol control policies—outlet density regulations among them—could be associated with lower odds of binge drinking among non-Hispanic/Latinx White but not non-Hispanic/Latinx Black or Hispanic/Latinx residents.

This study aims to investigate (1) whether ZIP code-level off-premise AOD is associated with binge drinking among a large sample of PWID who use opioids; (2) whether non-Hispanic/Latinx Black and Hispanic/Latinx PWID live in ZIP codes with higher AODs than their non-Hispanic/Latinx White counterparts; and (3) whether PWID race/ethnicity moderates the relationship between AOD and binge drinking.

## 2. METHODS

### 2.1 Overview

This study leverages data from the Centers for Disease Control and Prevention’s National HIV Behavioral Surveillance (NHBS) system, designed to monitor HIV-related behaviors of populations disproportionately burdened by HIV in large metropolitan statistical areas (MSAs) in the US. The NHBS study is the largest data source on health behaviors and outcomes among PWID and ensures a broad and racially/ethnically diverse sample of PWID in the metropolitan US. This multilevel cross-sectional analysis linked 2018 NHBS data on 9,660 PWID in 22 MSAs with ZIP code-level AOD data from the US Census Bureau’s 2016 ZIP Code Business Pattern (CBP) survey. Complete AOD data after 2016 was unavailable due to CBP’s suppression of cells for ZIP codes with fewer than three establishments. We thus linked the most recent and complete AOD data in 2016 with 2018 NHBS data (two-year lag) to investigate potential temporal relationships.

### 2.2 Sample

NHBS sought to recruit approximately 500 PWID from each of 23 MSAs in 2018 using respondent-driven sampling (RDS). Participants were eligible if they 1) reported injecting a non-prescribed drug in the past 12 months; 2) were aged ≥ 18; 3) lived in a participating MSA; and 4) could complete the survey in English or Spanish. This analysis further restricted the sample to participants who reported past-year injection or non-injection use of a non-prescribed opioid; 99% injected non-prescribed opioids. The analysis sample excluded participants who 1) reported a race/ethnicity other than Black, non-Hispanic/Latinx; Hispanic/Latinx; or White, non-Hispanic/Latinx due to sparse data, 2) had missing covariates or ZIP codes (< 5%), 3) did not report male or female sex due to the standard definition of binge drinking (5 or more drinks for males, 4 or more for females), and 4) lived in the San Juan-Bayamon MSA at the time of data collection because of a lack of racial/ethnic diversity. We did not include a measure of racial segregation because of possible multicollinearity between segregation and AOD that may obfuscate the independent effect of AOD on binge drinking.

### 2.3 Measures

#### 2.3.1 Outcome

*Past 30-day binge drinking.* Binge drinking is defined as consuming five or more drinks in one occasion for males and four or more for females (Centers for Disease Control and Prevention, 2022). The outcome was a dichotomous variable indicating whether the participant binge drank in the past 30 days, based on the question: “During the past 30 days, how many times did you drink 5 (4 if female) or more drinks of any kind of alcohol in about two hours?” *Primary independent variable*

#### 2.3.2 Exposure

*ZIP code-level alcohol outlet density:* NHBS queries the ZIP code of residence. This self-reported variable links individual participants to the 2016 AOD measure. The numerator for AOD was derived from the U.S. Census Bureau’s CBP data, and the denominator was the number of square miles in the ZIP code, approximated by Census ZIP code tabulation areas (United States Census Bureau, 2016). CBP reports annual, sub-national economic data on business establishments by industry type; these data are highly correlated with state and local administrative records (Matthews et al., 2011). Our measure was limited to off-premise outlets because regulating the density of off-premise outlets has been shown to be more effective in reducing harmful drinking than regulating on-premise outlets (i.e., bars and restaurants) (Campbell et al., 2009; Nesoff et al., 2021; Trangenstein et al., 2018). This measure’s numerator included supermarkets/grocery stores, convenience stores, beer/wine/liquor stores, gas stations with convenience stores, and pharmacies/drug stores that could sell alcohol for off-premise consumption per state liquor licensing laws.

#### 2.3.3 Covariates

##### State-level covariates

We used 2017 (one-year lag) *alcohol policy scores* quantifying the strength of states’ alcohol policy landscapes (Blanchette et al., 2020). Scores ranged from 0-100, with higher scores indicating more comprehensive and restrictive environments with regards to 21 policies governing alcohol production, sales, consumption, or furnishing practices (Naimi et al., 2014). We linked PWID to states using their reported county of residence.

##### ZIP code-level covariates

We used 2017 (one-year lag) United States Postal Services Address Information Systems data (United States Postal Service, 2017) to construct ZIP code-level *residential and business vacancies* by dividing the monthly average of vacancies by ZCTA square miles; the density of vacant buildings is associated with binge drinking and other harmful drinking patterns (Bernstein et al., 2007; Garvin et al., 2013). We also culled 2017 (one-year lag) ZIP code-level percent of people with income at or below the federal poverty line using the U.S. Census Bureau’s American Community Survey 5-Year data (United States Census Bureau, 2019).

##### Individual-level covariates

Participant data included sociodemographic characteristics (race/ethnicity, sex, age, income, education, employment), health insurance, incarceration (past 12 months), unhoused status (past 12 months), HIV serostatus, disability status using the DHHS data standard items (US Department of Health and Human Services Office of Minority Health, 2018;Brault et al., 2007), psychological distress as measured by the Kessler-6 scale (Kessler et al., 2003), network size, and respondent status as an RDS seed. We also included a covariate for receiving sterile needles from a syringe exchange program in the past year, as these programs often incorporate or refer participants to alcohol use prevention and treatment (Frost et al., 2018).

#### 2.3.4 Potential effect modifier

##### Individual race/ethnicity

We analyzed NHBS participants’ self-reported data to create three mutually exclusive racial/ethnic groups: Hispanic/Latinx; non-Hispanic/Latinx White (hereafter ‘White’); and non-Hispanic/Latinx Black (hereafter ‘Black’). When participants reported that they belonged to two racial groups and were not Hispanic/Latinx, we assigned them to a single racial/ethnic group using the Office of Management and Budget’s “plurality” guidelines (US Office of Management and Budget, 2016).

### 2.4 Analysis

Statistical analyses were conducted using SAS version 9.4. We explored distributions of all variables, examined correlations, and checked for multicollinearity (variance inflation factor ≤ 1.5 and tolerance ≥ 0.72 across all variables). We compared average AODs by PWID race/ethnicity using a Kruskal-Wallis test (McKight & Najab, 2010).

For multivariable analyses, we used three-level hierarchical generalized linear models with a logit link (Raudenbush & Bryk, 2002) to examine the relationship between ZIP code-level AOD and binge drinking. We modeled random intercepts for PWID clustered within ZIP codes and ZIP codes clustered within MSAs. Only three states contained more than one NHBS MSA so models could not support a random intercept for both MSA and state (unconditional models with MSAs and states resulted in a state-level variance component of zero [p > .05] and a warning that the estimated G matrix was not positive definite). Previous work has shown that when the highest level (here, state) of clustering is ignored in a model, that level’s variance component is redistributed to the lower level (Tranmer & Steel, 2001), so we modeled MSAs as the highest level of clustering in analysis.

We report three models: 1) an unadjusted model, 2) a multivariable model adjusted for ZIP code, state, and individual PWID covariates, and 3) an adjusted multivariable model with interactions to explore whether the relationship of ZIP code-level AOD to binge drinking varied by race/ethnicity. We present model results as odds ratios for binge drinking where the ZIP code-level AOD is in increments of three outlets per square mile above the median (i.e., approximately half of the interquartile range), allowing for a more intuitive understanding of how higher ZIP code-level AODs relate to the odds of binge drinking. We based increments on the median, as the sample distribution of AOD was highly positively skewed. To probe how the relationship between AOD and binge drinking depends on race/ethnicity in the interaction model, we 1) estimated simple slopes for the relationship between AOD and binge drinking for each race/ethnicity and tested differences in slopes and 2) estimated the predicted probabilities of binge drinking for each racial/ethnic group when the AOD is at the 25^th^ percentile, median, and 75^th^ percentile and conducted pairwise tests. Lastly, we plotted the predicted probabilities for each racial/ethnic group to visually compare differences in slopes across the entire range of ZIP code-level AODs (0-266.16).

### 2.5 Ethics

[BLINDED] Institutional Review Board approved all analyses.

## 3. RESULTS

### 3.1 Sample description

The sample included 9,660 PWID residing across 20 states and 22 MSAs. Approximately two-thirds (69%) were male; 45% were White, 38% were Black, and 18% were Hispanic/Latinx (Table 1). Most participants were experiencing poverty: 75% subsisted at or below the federal poverty guidelines, 85% were not employed or were unable to work due to health, and 67% had been unhoused in the past 12 months. More than a quarter (27%) reported binge drinking in the past 30 days, with prevalence rates of 32% for Black PWID, 28% for Hispanic/Latinx PWID, and 22% for White PWID (prevalence rates not shown in table).

**Table 1:** ZIP code, state and participant characteristics, overall and by past 30-day binge drinking (n=9,660)

|  | Overall<br>(n=9,660) | Binge drinking = No<br>(n=7,040; 72.88%)<br>Mean (SD) or N (%) | Binge drinking = Yes<br>(n=2,620; 27.12%) |
| --- | --- | --- | --- |
| **ZIP code (n=1,207 unique ZIP codes)** |  |  |  |
| Alcohol outlet density, mean (SD) | 11.54 (25) | - | - |
| Business vacancy density, mean (SD) | 64.10 (326) | - | - |
| Residential vacancy density, mean (SD) | 53.89 (116) | - | - |
| Percent of residents with income at or below the federal poverty line<sup>a</sup>, mean (SD) | 17.34 (10) | - | - |
| **State (n=20)** |  |  |  |
| Alcohol policy score, mean (SD) | 38.28 (9) | - | - |
| **Individual (NHBS)** |  |  |  |
| Binge drinking (past 30-days) | 2,620 (27) |  |  |
| Age, mean (SD) | 44.23 (13) | 44.03 (13) | 44.79 (12) |
| Race/ethnicity<sup>b</sup> |  |  |  |
| Black non-Hispanic/Latinx | 3,637 (38) | 2,464 (35) | 1,173 (45) |
| Hispanic/Latinx | 1,702 (18) | 1,218 (17) | 484 (18) |
| White non-Hispanic/Latinx | 4,321 (45) | 3,358 (48) | 963 (37) |
| Sex<sup>b</sup> |  |  |  |
| Male | 6,655 (69) | 4,847 (69) | 1,808 (69) |
| Female | 3,005 (31) | 2,193 (31) | 812 (31) |
| Income at or below the federal poverty level | 7,205 (75) | 5,199 (74) | 2,006 (77) |
| High-school graduate/General equivalency diploma | 6,942 (72) | 5,183 (74) | 1,759 (67) |
| Employment<sup>b</sup> |  |  |  |
| Not employed/other | 5,752 (60) | 4,186 (60) | 1,566 (60) |
| Employed full or part-time | 1,469 (15) | 1,049 (15) | 420 (16) |
| Unable to work due to health | 2,439 (25) | 1,805 (26) | 634 (24) |
| Daily injection | 8,564 (89) | 6,263 (89) | 2,301 (88) |
| HIV positive | 560 (6) | 420 (6) | 140 (5) |
| Incarceration (past 12-months) | 3,572 (37) | 2,585 (37) | 987 (38) |
| Unhoused (past 12-months) | 6,515 (67) | 4,645 (66) | 1,870 (72) |
| Insured | 7,237 (75) | 5,392 (77) | 1,845 (70) |
| Received clean needles from an SSP | 5,085 (53) | 3,948 (56) | 1,137 (43) |
| Psychological distress (past 30-days) | 3,722 (39) | 2,544 (36) | 1,178 (45) |
| Disability | 6,517 (67) | 4,593 (65) | 1,924 (73) |

<sup>a</sup>Data derived from the Census Bureau - American Community Survey, not individual NHBS data<sup>b</sup>Percentages may not add to 100% due to rounding

Among the 1,207 unique ZIP codes where PWID resided, the mean AOD was 11.5 (SD=25) and the median was 4.0 (range: 0-266), signifying a positively skewed distribution. The median AOD was 9.4 (SD=41) for Hispanic/Latinx PWID, 5.7 (SD=20) for White PWID, and 5.0 (SD=13) for Black PWID. Pairwise differences in median AOD were all statistically significant (p < 0.001).

### 3.2 Model-based analysis

In the unadjusted model, the relationship between ZIP-level AOD and binge drinking was null (OR: 1.01, 95% CI: 1.00-1.01, p = 0.12) (Table 2). In the adjusted model controlling for state-level alcohol policy scores and ZIP code- and individual-level covariates, there was a 2% increase in the odds of binge drinking for every three outlets per square mile—approximately half of the interquartile range—above the median (OR 1.02, 95% CI: 1.01-1.03, p = 0.002). In the model testing race/ethnicity as a moderator, simple slopes for Hispanic/Latinx (1.01) and White (1.03) PWID were statistically significant (p < 0.05) and showed that increases in AOD were associated with increases in binge drinking for these groups (Table 2). The simple slope for White PWID was significantly larger than that for Hispanic/Latinx PWID (p = 0.04).

**Table 2.**
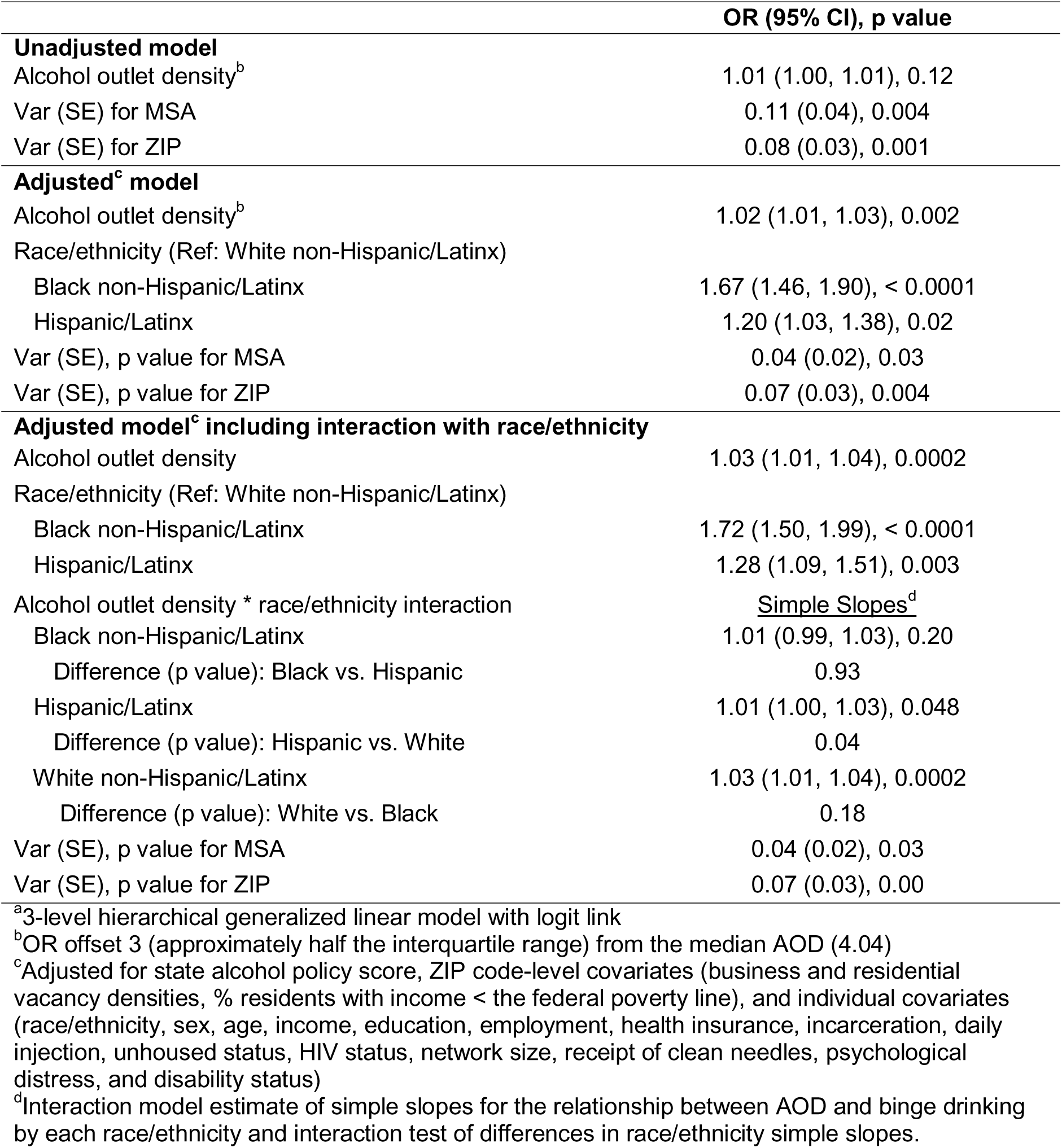
Unadjusted and adjusted hierarchical generalized linear regressions^a^ of the odds of past 30-day binge drinking on alcohol outlet density (AOD) and race/ethnicity potential effect modifier, 2018 (N=9,660)

To further probe the significant interaction between AOD and race/ethnicity, we estimated the predicted probability of binge drinking, for each race/ethnicity, across a range of AODs (Table 3). At the median AOD, predicted probabilities were significant for all races, but the magnitude varied by race/ethnicity (Table 3). Specifically, the predicted probability of binge drinking at the median AOD was highest for Black (0.28, 95% CI: 0.24-0.33, p < 0.001) and lowest for White PWID (0.18, 95% CI: 0.15-0.22, p < 0.001). All pairwise differences were significant at p < 0.05 (Table 3). Predicted probabilities were nearly identical (p < 0.001) to those at the median when the AOD was set to the 25^th^ (1.8) and 75^th^ (8.5) percentiles for unique ZIP codes with significant pairwise differences (p < 0.05). However, differences between racial/ethnic groups were no longer significant for White vs. Hispanic/Latinx PWID at 25 or more outlets per square mile, White vs. Black PWID at 55 or more outlets per square mile, and Hispanic/Latinx vs. Black PWID at 60 or more outlets per square mile. Figure 1 plots predicted probabilities of binge drinking by race/ethnicity across the range of AODs.

**Table 3.** Race interaction model^a, b^: Predicted probabilities for past 30-day binge drinking by race/ethnicity, 2018 (N=9,660)

|  | Predicted probability<sup>c</sup> of binge drinking<br>Probability (95% CI), p value | Pairwise differences<br>p value |
| --- | --- | --- |
| Non-Hispanic Black | 0.28 (0.24, 0.33), <.0001 | Non-Hispanic Black vs. White<br>< 0.0001 |
| Hispanic | 0.23 (0.18, 0.27), <.0001 | Hispanic vs. Non-Hispanic Black<br>0.001 |
| Non-Hispanic White | 0.18 (0.15, 0.22), <.0001 | Non-Hispanic White vs. Hispanic<br>0.003 |

<sup>a</sup>3-level (Level 1: Individual PWID, Level 2: ZIP code, Level 3: MSA) hierarchical generalized linear model, with logit link
<sup>b</sup>Alcohol outlet density \* race/ethnicity interaction model adjusted for state-level alcohol policy score, ZIP code-level covariates (density of business and residential vacancies, percent of residents with income at or below the federal poverty line), and individual covariates (race/ethnicity, sex, age, income, education, employment, health insurance, incarceration, daily injection status, unhoused status, HIV status, network size, receipt of clean needles from an SSP, psychological distress, and disability status)
<sup>c</sup>Predicted probability and pairwise differences when alcohol outlet density set to the median (4.04) for unique ZIP codes

As a sensitivity analysis, we ran models excluding alcohol policy scores. The relationship between AOD and binge drinking was unchanged (no or negligible change to the estimates and confidence intervals).

## 4. DISCUSSION

This study investigated the relationship between ZIP code-level off-premise AOD and binge drinking among PWID who reported non-prescribed opioid use, overall and by race/ethnicity. The median AOD across ZIP codes was 4.0 outlets per square mile. For every increase of three outlets per square mile above the median, the odds of binge drinking overall increased by 2%. However, exposure to alcohol outlets and its association with binge drinking varied by race/ethnicity. Hispanic/Latinx PWID lived in ZIP codes with a significantly higher median AOD compared to the other racial/ethnic groups. Additionally, increases in AOD were significantly associated with increases in binge drinking among Hispanic/Latinx and White PWID but not among Black PWID.

There was a high prevalence of binge drinking among the participants, with over one in four reporting binge drinking in the past 30 days; in contrast, the prevalence of binge drinking among the general US adult population is approximately 17% (Bohm et al., 2021). The elevated prevalence among our large PWID sample aligns with other US-based studies, where past-30-day prevalences for binge drinking among PWID have ranged from 19% to 40% (Earlywine et al., 2021; Marcus et al., 2020; Park et al., 2022; Rushmore et al., 2023; Wise et al., 2023; Wu et al., 2022). This high prevalence of binge drinking among PWID is notable. Compared to those who use opioids through other administration routes, the risk of overdose is higher among people who inject opioids due to the immediate and intense effects of injected drugs (Mathers et al., 2013). Binge drinking may amplify this risk in this vulnerable population by exacerbating opioids’ suppressive effects on respiration.

Importantly, Black and Hispanic/Latinx PWID had higher prevalences of binge drinking than their White counterparts, and also significantly higher probabilities of binge drinking at the median AOD among unique ZIP codes. While no other studies have explored racial/ethnic differences in drinking behaviors among PWID, the 2023 National Survey on Drug Use and Health similarly found that Hispanic/Latinx adults in the general population had a higher prevalence of binge drinking than White adults (25.7 vs. 23.9%), though the prevalence of binge drinking was similar among Black and White adults (23.8% vs. 23.9%) (National Institute on Alcohol Abuse and Alcoholism, 2024). Given that Black PWID in this study reported a higher prevalence of binge drinking, researchers may explore binge drinking as a key intervention target to reverse surging overdose rates among Black PWID and others who use opioids (Friedman & Hansen, 2022; Han et al., 2022; Kariisa et al., 2022).

Parallel to findings for the general population, in this sample of PWID Hispanic/Latinx participants tend to live in ZIP codes with higher AODs compared to White participants (Berke et al., 2010; Fliss et al., 2021; LaVeist & Wallace, 2000; Lee et al., 2020; Romley et al., 2007). One potential explanatory mechanism may be that Hispanic/Latinx PWID reside in ZIP codes targeted for alcohol outlets due to evolving perceived market opportunities. Liquor companies, recognizing the increasing share of Hispanic/Latinx people of legal drinking age, are increasingly focusing on marketing alcohol to these consumers, a pattern evident in recent industry trends (Chaudhuri, 2017). This focus may drive higher AOD in ZIP codes with significant Hispanic/Latinx PWID. In our study, White and Black PWID tend to live in ZIP codes with similar densities of off-premise outlets.

On initial examination, our findings indicate that residing in ZIP codes with higher than the median AOD is associated with increased odds of binge drinking for our entire sample of PWID. This finding aligns with previous research highlighting the impact of AOD on alcohol consumption among the US urban general population (Auchincloss et al., 2022; Ahern et al., 2013). However, the race/ethnicity interaction model demonstrated that this association is only significant among Hispanic/Latinx and White PWID, with a stronger effect observed among White PWID. This suggests that policies limiting AOD may be effective in reducing binge drinking among White PWID and, to a lesser extent, Hispanic/Latinx PWID. Evidence from the general population supports the effectiveness of AOD regulation in curbing binge drinking through zoning and licensing policies that reduce alcohol outlet density and subsequently restrict alcohol availability (Bryden et al., 2012; Campbell et al., 2009; Popova et al., 2009; Xuan et al., 2015). By targeting AOD in areas with higher densities, these interventions could potentially mitigate some of the alcohol-related harms experienced by White and Hispanic/Latinx PWID.

Notably, however, the interplays found here among individual race/ethnicity, ZIP code-level AOD, and binge drinking raises the need for equitable prevention strategies and a corollary caution about uniform approaches to AOD regulation potentially exacerbating inequities. While Black PWID had a higher predicted probability of binge drinking compared to White and Hispanic/Latinx PWID in ZIP codes with AODs close to the median, this difference became non-significant in ZIP codes with the highest outlet densities (55 outlets per square mile or greater for White vs. Black predicted probability comparison). This may indicate that regulations reducing ZIP code-level AODs toward the overall median curbs binge drinking most among White PWID and least among their Black counterparts. Thus, AOD regulation may be inadequate for addressing the needs of Black PWID, and prevention efforts should prioritize approaches that consider the unique needs of this population. These strategies must be co-created with local communities and coupled with equitable implementation to address alcohol-related harms without inadvertently exacerbating others.

Certain methodological limitations warrant consideration. First, the use of 2018 NHBS data and 2016 AOD data, the most recent available at the time of analysis, may obscure contemporary trends in alcohol availability and binge drinking, respectively. The proliferation of home delivery and alcohol to-go services during the COVID-19 pandemic has expanded the traditional definition of off-premise access, increasing local alcohol availability (Haley, Peddireddy, et al., 2023). Binge drinking among the general population also increased during the pandemic (Barbosa et al., 2021). Given these shifts, our estimates for outlet density and binge drinking may be conservative. Recent studies have demonstrated a greater magnitude of the effect of off-premise alcohol availability on alcohol consumption post-pandemic (Grossman et al., 2022; Trangenstein et al., 2023). As a result, our estimate for the association between off-premise outlet density and binge drinking may also be conservative. Similarly, CBP data suppress outlets with fewer than ten employees, only capture registered or licensed businesses, and might misclassify some alcohol outlets, leading to potential underreporting of ZIP code-level AOD. Also, 67% of the sample were unhoused at some point during the previous year, and their reported residential ZIP codes may not correspond with the locations where they purchased and consumed alcohol. The survey also did not assess whether participants used alcohol and opioids simultaneously. Therefore, we could not investigate the association of co-use with binge drinking or other harms such as overdose. Findings are not generalizable to all PWID in the US, as participants were recruited from major MSAs only. Additionally, our analytic sample may not represent the broader populations of PWID within the sampled MSAs.

In this study, we highlighted the complex relationships among the alcohol risk environment, binge drinking, and individual race/ethnicity. Notably, reducing AOD may decrease binge drinking among White and Hispanic/Latinx PWID, but not among Black PWID. Factors beyond AOD may drive binge drinking behaviors among Black PWID, underscoring the need for more research on effective interventions for this subpopulation. Future research should explore these dynamics, using longitudinal designs to clarify causal relationships between changes in the alcohol risk environment and binge drinking. Additionally, mixed-methods approaches can provide deeper insights into how PWID engage with their alcohol environments. Finally, research should investigate the downstream effects of binge drinking among PWID, including its contribution to fatal and nonfatal drug overdoses.

## Data Availability

All data produced and analyzed in the present study are only available upon request to the Centers for Disease Control and Prevention's National HIV Behavioral Surveillance program.

## ACKNOWLEDGEMENTS

We express deep appreciation to the reviewers and Editor at *Drug and Alcohol Dependence* and our funders (R01DA046197, Cooper PI; K01DA051696, Yarbrough; K01DA046307, Haley; 5T32DA050552). The results and opinions expressed therein represent those of the authors and do not necessarily reflect those of NIH or NIDA. The funders had no role in relation to the study design; collection, analysis and interpretation of data; writing of the report; and decision to submit the article for publication. We are rateful to Jason Blanchette, JD, MPH (Boston University School of Public Health) and Tim Naimi, MD, MPH (University of Victoria) for providing the alcohol policy scores used in this analysis. Lastly, we would like to express our gratitude to the NHBS PWID participants and to the Principal Investigators and teams at the NHBS sites, including: Atlanta, GA: Pascale Wortley, Jeff Todd, David Melton; Baltimore, MD: Colin Flynn, Danielle German; Boston, MA: Monina Klevens, Rose Doherty, Conall O’Cleirigh; Chicago, IL: Antonio D. Jimenez, Thomas Clyde; Dallas, TX: Jonathon Poe, Margaret Vaaler, Jie Deng; Denver, CO: Alia Al-Tayyib, Daniel Shodell; Detroit, MI: Emily Higgins, Vivian Griffin, Corrine Sanger; Houston, TX: Salma Khuwaja, Zaida Lopez, Paige Padgett; Los Angeles, CA: Ekow Kwa Sey, Yingbo Ma, Hugo Santacruz; Memphis, TN: Meredith Brantley, Christopher Mathews, Jack Marr; Miami, FL: Emma Spencer, Willie Nixon, David Forrest; Nassau-Suffolk, NY: Bridget Anderson, Ashley Tate, Meaghan Abrego; New Orleans, LA: William T. Robinson, Narquis Barak, Jeremy M. Beckford; New York City, NY: Sarah Braunstein, Alexis Rivera, Sidney Carrillo Newark, NJ: Abdel R. Ibrahim, Afework Wogayehu, Luis Moraga; Philadelphia, PA: Kathleen A. Brady, Jennifer Shinefeld, Chrysanthus Nnumolu,; Portland, OR: Timothy W. Menza, E. Roberto Orellana, Amisha Bhattari; San Diego, CA: Anna Flynn, Onika Chambers, Marisa Ramos; San Francisco, CA: Willi McFarland, Jessica Lin, Desmond Miller; San Juan, PR: Sandra Miranda De León, Yadira Rolón-Colón, María Pabón Martínez; Seattle, WA: Tom Jaenicke, Sara Glick; Virginia Beach, VA: Jennifer Kienzle, Brandie Smith, Toyah Reid; Washington, DC: Jenevieve Opoku, Irene Kuo.

## REFERENCES

Ahern, J., Margerison-Zilko, C., Hubbard, A., & Galea, S. (2013). Alcohol outlets and binge drinking in urban neighborhoods: the implications of nonlinearity for intervention and policy. American Journal of Public Health, 103(4), e81–7. 10.2105/AJPH.2012.301203

Arasteh, K., Des Jarlais, D. C., & Perlis, T. E. (2008). Alcohol and HIV sexual risk behaviors among injection drug users. Drug and Alcohol Dependence, 95(1–2), 54–61. 10.1016/j.drugalcdep.2007.12.008

Auchincloss, A. H., Niamatullah, S., Adams, M., Melly, S. J., Li, J., & Lazo, M. (2022). Alcohol outlets and alcohol consumption in changing environments: prevalence and changes over time. *Substance Abuse Treatment*, Prevention, and Policy, 17(1), 7. 10.1186/s13011-021-00430-6

Barbosa, C., Cowell, A. J., & Dowd, W. N. (2021). Alcohol Consumption in Response to the COVID-19 Pandemic in the United States. Journal of Addiction Medicine, 15(4), 341–344. 10.1097/ADM.0000000000000767

Berke, E. M., Tanski, S. E., Demidenko, E., Alford-Teaster, J., Shi, X., & Sargent, J. D. (2010). Alcohol retail density and demographic predictors of health disparities: a geographic analysis. American Journal of Public Health, 100(10), 1967–1971. 10.2105/AJPH.2009.170464

Bernstein, K. T., Galea, S., Ahern, J., Tracy, M., & Vlahov, D. (2007). The built environment and alcohol consumption in urban neighborhoods. Drug and Alcohol Dependence, 91(2–3), 244–252. 10.1016/j.drugalcdep.2007.06.006

Blanchette, J. G., Lira, M. C., Heeren, T. C., & Naimi, T. S. (2020). Alcohol Policies in U.S. States, 1999-2018. Journal of Studies on Alcohol and Drugs, 81(1), 58–67. 10.15288/jsad.2020.81.58

Bohm, M. K., Liu, Y., Esser, M. B., Mesnick, J. B., Lu, H., Pan, Y., & Greenlund, K. J. (2021). Binge drinking among adults, by select characteristics and state — United States, 2018. American Journal of Transplantation, 21(12), 4084–4091. 10.1111/ajt.16057

Brault, M., Stern, S., & Raglin, D. (2007). Evaluation Report Covering Disability. 2006 American Community Survey Content Test Report. . US Department of Health and Human Services.

Bryden, A., Roberts, B., McKee, M., & Petticrew, M. (2012). A systematic review of the influence on alcohol use of community level availability and marketing of alcohol. Health & Place, 18(2), 349–357. 10.1016/j.healthplace.2011.11.003

Campbell, C. A., Hahn, R. A., Elder, R., Brewer, R., Chattopadhyay, S., Fielding, J., Naimi, T. S., Toomey, T., Lawrence, B., Middleton, J. C., & Task Force on Community Preventive Services. (2009). The effectiveness of limiting alcohol outlet density as a means of reducing excessive alcohol consumption and alcohol-related harms. American Journal of Preventive Medicine, 37(6), 556–569. 10.1016/j.amepre.2009.09.028

Centers for Disease Control and Prevention. (2022, July 11). Excessive Alcohol Use. https://www.cdc.gov/chronicdisease/resources/publications/factsheets/alcohol.htm

Chaudhuri, S. (2017, May 5). Liquor Makers Step Up Efforts to Win Over Hispanic Drinkers. Wall Street Journal.

Earlywine, J. J., Bazzi, A. R., Biello, K. B., & Klevens, R. M. (2021). High Prevalence of Indications for Pre-exposure Prophylaxis Among People Who Inject Drugs in Boston, Massachusetts. American Journal of Preventive Medicine, 60(3), 369–378. 10.1016/j.amepre.2020.09.011

Esser, M. B., Pickens, C. M., Guy, G. P., & Evans, M. E. (2021). Binge Drinking, Other Substance Use, and Concurrent Use in the U.S., 2016-2018. American Journal of Preventive Medicine, 60(2), 169–178. 10.1016/j.amepre.2020.08.025

Fliss, M. D., Cox, M. E., Wallace, J. W., Simon, M. C., Knuth, K. B., & Proescholdbell, S. (2021). Measuring and mapping alcohol outlet environment density, clusters, and racial and ethnic disparities in durham, north carolina, 2017. Preventing Chronic Disease, 18, E89. 10.5888/pcd18.210127

Fournier, C., Ghabrash, M. F., Artenie, A., Roy, E., Zang, G., Bruneau, J., & Jutras-Aswad, D. (2018). Association between binge drug use and suicide attempt among people who inject drugs. Substance AbuselJ: Official Publication of the Association for Medical Education and Research in Substance Abuse, 39(3), 315–321. 10.1080/08897077.2017.1389800

Friedman, J. R., & Hansen, H. (2022). Evaluation of Increases in Drug Overdose Mortality Rates in the US by Race and Ethnicity Before and During the COVID-19 Pandemic. JAMA Psychiatry, 79(4), 379–381. 10.1001/jamapsychiatry.2022.0004

Frost, M. C., Williams, E. C., Kingston, S., & Banta-Green, C. J. (2018). Interest in getting help to reduce or stop substance use among syringe exchange clients who use opioids. Journal of Addiction Medicine, 12(6), 428–434. 10.1097/ADM.0000000000000426

Garvin, E., Branas, C., Keddem, S., Sellman, J., & Cannuscio, C. (2013). More than just an eyesore: local insights and solutions on vacant land and urban health. Journal of Urban Health, 90(3), 412–426. 10.1007/s11524-012-9782-7

Grossman, E. R., Benjamin-Neelon, S. E., & Sonnenschein, S. (2022). Alcohol consumption and alcohol home delivery laws during the COVID-19 pandemic. Substance AbuselJ: Official Publication of the Association for Medical Education and Research in Substance Abuse, 43(1), 1139–1144. 10.1080/08897077.2022.2060432

Haley, S. J., Jardine, S. J., Kelvin, E. A., Herrmann, C., & Maroko, A. R. (2023). Neighborhood Alcohol Outlet Density, Historical Redlining, and Violent Crime in NYC 2014-2018. International Journal of Environmental Research and Public Health, 20(4). 10.3390/ijerph20043212

Haley, S. J., Peddireddy, S., El-Harakeh, A., Akasreku, B., & Riibe, D. (2023). Qualitative study of states’ capacity to support alcohol prevention policies during the COVID-19 pandemic in the USA. Drug and Alcohol Review, 42(6), 1358–1374. 10.1111/dar.13714

Han, B., Einstein, E. B., Jones, C. M., Cotto, J., Compton, W. M., & Volkow, N. D. (2022). Racial and Ethnic Disparities in Drug Overdose Deaths in the US During the COVID-19 Pandemic. JAMA Network Open, 5(9), e2232314. 10.1001/jamanetworkopen.2022.32314

Irvin, R., Chander, G., Falade-Nwulia, O., Astemborski, J., Starbird, L., Kirk, G. D., Sulkowski, M. S., Thomas, D. L., & Mehta, S. H. (2019). Overlapping epidemics of alcohol and illicit drug use among HCV-infected persons who inject drugs. Addictive Behaviors, 96, 56–61. 10.1016/j.addbeh.2019.04.023

Kariisa, M., Davis, N. L., Kumar, S., Seth, P., Mattson, C. L., Chowdhury, F., & Jones, C. M. (2022). Vital Signs: Drug Overdose Deaths, by Selected Sociodemographic and Social Determinants of Health Characteristics - 25 States and the District of Columbia, 2019-2020. MMWR. Morbidity and Mortality Weekly Report, 71(29), 940–947. 10.15585/mmwr.mm7129e2

Kessler, R. C., Barker, P. R., Colpe, L. J., Epstein, J. F., Gfroerer, J. C., Hiripi, E., Howes, M. J., Normand, S.-L. T., Manderscheid, R. W., Walters, E. E., & Zaslavsky, A. M. (2003). Screening for serious mental illness in the general population. Archives of General Psychiatry, 60(2), 184–189. 10.1001/archpsyc.60.2.184

LaVeist, T. A., & Wallace, J. M. (2000). Health risk and inequitable distribution of liquor stores in African American neighborhood. Social Science & Medicine, 51(4), 613–617. 10.1016/s0277-9536(00)00004-6

Lee, J. P., Ponicki, W., Mair, C., Gruenewald, P., & Ghanem, L. (2020). What explains the concentration of off-premise alcohol outlets in Black neighborhoods? SSM - Population Health, 12, 100669. 10.1016/j.ssmph.2020.100669

Lynch, E. E., Malcoe, L. H., Laurent, S. E., Richardson, J., Mitchell, B. C., & Meier, H. C. S. (2021). The legacy of structural racism: Associations between historic redlining, current mortgage lending, and health. SSM - Population Health, 14, 100793. 10.1016/j.ssmph.2021.100793

Marcus, R., Cha, S., Sionean, C., Kanny, D., & National HIV Behavioral Surveillance Study Group. (2020). HIV Injection Risk Behaviors among HIV-Negative People Who Inject Drugs Experiencing Homelessness, 23 U.S. Cities. Journal of Social Distress and the Homeless, 1(9). 10.1080/10530789.2021.1892931

Mathers, B. M., Degenhardt, L., Bucello, C., Lemon, J., Wiessing, L., & Hickman, M. (2013). Mortality among people who inject drugs: a systematic review and meta-analysis. Bulletin of the World Health Organization, 91(2), 102–123. 10.2471/BLT.12.108282

Matthews, S. A., McCarthy, J. D., & Rafail, P. S. (2011). Using ZIP code business patterns data to measure alcohol outlet density. Addictive Behaviors, 36(7), 777–780. 10.1016/j.addbeh.2011.02.009

McKight, P. E., & Najab, J. (2010). Kruskal-Wallis Test. In I. B. Weiner & W. E. Craighead (Eds.), The corsini encyclopedia of psychology. John Wiley & Sons, Inc. 10.1002/9780470479216.corpsy0491

Naimi, T. S., Blanchette, J., Nelson, T. F., Nguyen, T., Oussayef, N., Heeren, T. C., Gruenewald, P., Mosher, J., & Xuan, Z. (2014). A new scale of the U.S. alcohol policy environment and its relationship to binge drinking. American Journal of Preventive Medicine, 46(1), 10–16. 10.1016/j.amepre.2013.07.015

National Institute on Alcohol Abuse and Alcoholism. (2024). Alcohol Use in the United States: Age Groups and Demographic Characteristics. https://www.niaaa.nih.gov/alcohols-effects-health/alcohol-topics/alcohol-facts-and-statistics/alcohol-use-united-states-age-groups-and-demographic-characteristics

Nesoff, E. D., Milam, A. J., Morrison, C., Weir, B. W., Branas, C. C., Furr-Holden, D. M., Knowlton, A. R., & Martins, S. S. (2021). Alcohol outlets, drug paraphernalia sales, and neighborhood drug overdose. The International Journal on Drug Policy, 95, 103289. 10.1016/j.drugpo.2021.103289

Park, D., Oh, S., Cano, M., Salas-Wright, C. P., & Vaughn, M. G. (2022). Trends and distinct profiles of persons who inject drugs in the United States, 2015-2019. Preventive Medicine, 164, 107289. 10.1016/j.ypmed.2022.107289

Popova, S., Giesbrecht, N., Bekmuradov, D., & Patra, J. (2009). Hours and days of sale and density of alcohol outlets: impacts on alcohol consumption and damage: a systematic review. Alcohol and Alcoholism, 44(5), 500–516. 10.1093/alcalc/agp054

Raudenbush, S. W., & Bryk, A. S. (2002). Hierarchical Linear Models: Applications and Data Analysis Methods (illustrated ed.). SAGE.

Robert Wood Johnson Foundation, & University of Wisconsin Population Health Institute. (n.d.). Strategies | County Health Rankings & Roadmaps. Retrieved June 15, 2023, from https://www.countyhealthrankings.org/take-action-to-improve-health/what-works-for-health/strategies

Romley, J. A., Cohen, D., Ringel, J., & Sturm, R. (2007). Alcohol and environmental justice: the density of liquor stores and bars in urban neighborhoods in the United States. Journal of Studies on Alcohol and Drugs, 68(1), 48–55. 10.15288/jsad.2007.68.48

Rushmore, J., Buchacz, K., Broz, D., Agnew-Brune, C. B., Jones, M. L. J., Cha, S., & NHBS Study Group. (2023). Factors Associated with Exchange Sex Among Cisgender Persons Who Inject Drugs: Women and MSM-23 U.S. Cities, 2018. AIDS and Behavior, 27(1), 51–64. 10.1007/s10461-022-03743-0

Sacks, J. J., Brewer, R. D., Mesnick, J., Holt, J. B., Zhang, X., Kanny, D., Elder, R., & Gruenewald, P. J. (2020). Measuring alcohol outlet density: an overview of strategies for public health practitioners. Journal of Public Health Management and PracticelJ: JPHMP, 26(5), 481–488. 10.1097/PHH.0000000000001023

Substance Abuse and Mental Health Services Administration. (2022). Implementing Community-Level Policies to Prevent Alcohol Misuse. SAMHSA.

Tori, M. E., Larochelle, M. R., & Naimi, T. S. (2020). Alcohol or Benzodiazepine Co-involvement With Opioid Overdose Deaths in the United States, 1999-2017. JAMA Network Open, 3(4), e202361. 10.1001/jamanetworkopen.2020.2361

Trangenstein, P. J., Curriero, F. C., Webster, D., Jennings, J. M., Latkin, C., Eck, R., & Jernigan, D. H. (2018). Outlet type, access to alcohol, and violent crime. Alcoholism, Clinical and Experimental Research, 42(11), 2234–2245. 10.1111/acer.13880

Trangenstein, P. J., Gray, C., Rossheim, M. E., Sadler, R., & Jernigan, D. H. (2020). Alcohol outlet clusters and population disparities. Journal of Urban Health, 97(1), 123–136. 10.1007/s11524-019-00372-2

Trangenstein, P. J., Greenfield, T. K., Karriker-Jaffe, K. J., & Kerr, W. C. (2023). Beverage- and context-specific alcohol consumption during COVID-19 in the United States: The role of alcohol to-go and delivery purchases. Journal of Studies on Alcohol and Drugs. 10.15288/jsad.22-00408

Tranmer, M., & Steel, D. G. (2001). Ignoring a Level in a Multilevel Model: Evidence from UK Census Data. Environment & Planning A, 33(5), 941–948. 10.1068/a3317

United States Census Bureau. (2016). 2016 U.S Census Bureau’s ZIP Codes Business Pattern datasets. U.S. Census Bureau Survey Office. https://www.census.gov/data/developers/data-sets/cbp-nonemp-zbp/zbp-api.html

United States Census Bureau. (2019). American Community Survey 5-year estimates (2015-2019) Data Table S1701. . United States Census Bureau American Community Survey. https://www.census.gov/programs-surveys/acs/data.html

United States Postal Service. (2017). United States Postal Service Address Information Systems Data . https://postalpro.usps.com/address-quality/ais-viewer

US Department of Health and Human Services Office of Minority Health. (2018). Data Collection Standards for Race, Ethnicity, Primary Language, Sex, and Disability Status. https://minorityhealth.hhs.gov/data-collection-standards-race-ethnicity-primary-language-sex-and-disability-status

US Office of Management and Budget. (2016). Standards for Maintaining, Collecting, and Presenting Federal Data on Race and Ethnicity. US Office of Management and Budget.

van der Schrier, R., Roozekrans, M., Olofsen, E., Aarts, L., van Velzen, M., de Jong, M., Dahan, A., & Niesters, M. (2017). Influence of Ethanol on Oxycodone-induced Respiratory Depression: A Dose-escalating Study in Young and Elderly Individuals. Anesthesiology, 126(3), 534–542. 10.1097/ALN.0000000000001505

Wise, A., Kianian, B., Chang, H. H., Linton, S., Wolfe, M. E., Smith, J., Tempalski, B., Des Jarlais, D., Ross, Z., Semaan, S., Wejnert, C., Sionean, C., Cooper, H. L. F., & NHBS Study Group. (2023). Socioeconomic and racial/ethnic spatial polarization and incarceration among people who inject drugs in 19 US metropolitan areas, 2015. SSM - Population Health, 23, 101486. 10.1016/j.ssmph.2023.101486

Wu, K., Tie, Y., Dasgupta, S., Beer, L., & Marcus, R. (2022). Injection and Non-Injection Drug Use Among Adults with Diagnosed HIV in the United States, 2015-2018. AIDS and Behavior, 26(4), 1026–1038. 10.1007/s10461-021-03457-9

Xuan, Z., Blanchette, J., Nelson, T. F., Heeren, T., Oussayef, N., & Naimi, T. S. (2015). The alcohol policy environment and policy subgroups as predictors of binge drinking measures among US adults. American Journal of Public Health, 105(4), 816–822. 10.2105/AJPH.2014.302112

